# White Matter Microstructural Alteration in Type 2 Diabetes: A Combined UK Biobank Study of Diffusion Tensor Imaging and Neurite Orientation Dispersion and Density Imaging

**DOI:** 10.1101/2022.07.09.22277453

**Authors:** Abdulmajeed Alotaibi, Anna Podlasek, Amjad AlTokhis, Chris R. Tench, Ali-Reza Mohammadi-Nejad, Stamatios N. Sotiropoulos, Cris S. Constantinescu, Sieun Lee, Rob A. Dineen

## Abstract

**Background:** Type 2 diabetes mellitus impacts the brain microstructural environment. Diffusion tensor imaging (DTI) has been widely used to characterize white matter microstructural abnormalities in type 2 diabetes but fails to fully characterise disease effects on complex white matter tracts. Neurite orientation dispersion and density imaging (NODDI) has been proposed as an alternative to DTI with higher specificity to characterize white matter microstructures. Although NODDI has not been widely applied in diabetes, this biophysical model has the potential to investigate microstructural changes in white matter pathology.

**Aims and objectives:** (1) To investigate brain white matter alterations in people with type 2 diabetes using DTI and NODDI; (2) To assess the association between white matter changes in type 2 diabetes with disease duration and diabetes control as reflected by glycated haemoglobin (HbA1c) levels.

**Methods:** We examined white matter microstructure in 48 white matter tracts using data from the UK Biobank in 3,338 participants with type 2 diabetes (36% women, mean age 66 years) and 30,329 participants without type 2 diabetes (53% women, mean age 64 years). The participants had undergone 3.0T multiparametric brain imaging, including T1 weighted imaging and diffusion imaging for DTI and NODDI. Region of interest analysis of fractional anisotropy (FA), mean diffusivity (MD), axial diffusivity (AD), radial diffusivity (RD), orientation dispersion index (ODI), intracellular volume fraction (ICVF), and isotropic water fraction (IsoVF) were conducted to assess white matter abnormalities. A general linear model was applied to evaluate intergroup white matter differences and their association with the metabolic profile.

**Result:** Reduced FA and ICVF and increased MD, AD, RD, ODI, and IsoVF values were observed in participants with type 2 diabetes compared to non-type 2 diabetes participants (*P*<0.05). Reduced FA and ICVF in most white matter tracts were associated with longer disease duration and higher levels of HbA1c (0< r ≤0.2, *P*<0.05). Increased MD, AD, RD, ODI and IsoVF also correlated with longer disease duration and higher HbA1c (0< r ≤0.2, *P*<0.05).

**Discussion:** NODDI detected microstructural changes in brain white matter in participants with type 2 diabetes. The revealed abnormalities are proxies for lower neurite density and loss of fibre orientation coherence, which correlated with longer disease duration and an index of poorly controlled blood sugar. NODDI contributed to DTI in capturing white matter differences in participants with type 2 diabetes, suggesting the feasibility of NODDI in detecting white matter alterations in type 2 diabetes.

**Conclusion:** Type 2 diabetes can cause white matter microstructural abnormalities that have associations with glucose control. The NODDI diffusion model allows the characterisation of white matter neuroaxonal pathology in type 2 diabetes, giving biophysical information for understanding the impact of type 2 diabetes on brain microstructure.

## Introduction

Type 2 diabetes mellitus (T2DM) is a common metabolic disorder with a rising global prevalence^1^. It impacted approximately 462 million people in 2017, accounting for 6.28% of the worldwide population (4.4% of those aged 15-49 years, 15% of those aged 50-69 years, and 22% of those aged 70+), or a prevalence rate of 6059 cases per 100,000^2^. T2DM results in microvascular and macrovascular complications^3^, leading to microvascular impairments, generally recognised as risk factors for general brain atrophy, microstructural alterations, and cognitive impairment^4–6^. Using advanced neuroimaging techniques, researchers have recently focused on understanding the link between T2DM and microstructural abnormalities in the brain. Advanced magnetic resonance imaging (MRI) techniques such as Diffusion tensor imaging (DTI) have been widely used to explore microstructural changes in the brain, and it has been adopted in several investigations to identify global/local network abnormalities and microstructural changes in WM^7–9^. WM tracts microstructurally affected by T2DM as detected by DTI include the corpus callosum, corona radiata, cingulum, internal/external capsules, fornix, uncinate fasciculus, and corticospinal tracts^4,10,11^. DTI is based on a single compartment Gaussian diffusion model of brain microstructure with anisotropic water diffusion^12^. Although DTI is sensitive to WM microstructural changes, it lacks specificity for the tissue microstructural features of T2DM^13–15^. When two or more distinct tissues with heterogeneous diffusion properties are present in a single voxel, the interpretation of DTI values becomes more complicated^15^. Furthermore, DTI may not be an appropriate diffusion model to describe the non-Gaussian diffusion processes in biological tissues^16^.

An alternative biophysical diffusion model, known as neurite orientation dispersion and density imaging (NODDI), has been proposed to theoretically provides more specific characterisations of WM microstructure^17^. NODDI uses a multi-shell diffusion model with a high angular resolution that describes brain tissue as a simplified combination of three compartments and separates the signals from each compartment^17^. A Watson distribution assumption of sticks models the first compartment, known as the intracellular or intra-neurite compartment (axons and dendrites), and the signal of each stick is assumed to be a perfectly anisotropic diffusion tensor with a perpendicular diffusivity equal to zero. Also, it represents extracellular space with a Gaussian anisotropic diffusion and a free water compartment that represents an isotropic Gaussian diffusion, such as cerebrospinal fluids (CSF)^17^. NODDI uses three scalar parameters: neurite density index (NDI), also known as intra-cellular volume fraction (ICVF), orientation dispersion index (ODI) (0 for perfectly aligned straight fibres and 1 for entirely isotropic), and free water fraction or isotropic volume fraction (IsoVF)^18^. These parameters explicitly estimate the orientation dispersion and neurite density, contributing to conventional DTI parameters such as fractional anisotropy (FA)^17^.

Although NODDI has not been widely applied in diabetes, it is promising in evaluating the complexity of WM neuroaxonal pathology. A previous study has demonstrated that NODDI parameters can detect differences between T2DM patients and non-T2DM participants, suggesting their promising role as a biomarker for assessing WM microstructural deficits in T2DM patients^19^. However, this study was performed on 18 patients with T2DM only, the slice thickness was large, and the diffusion encoding direction of 25 was relatively small to estimate the fibre orientation. These technical and sample size limitations may have limited the quantification of NODDI-derived parameters in T2DM patients.

Herein, we explore the brain WM microstructural changes in a large cohort of participants with T2DM from a population-level study (the UK Biobank^20,21^) using measures derived from DTI and NODDI. The aims of this large-scale study are 1) to investigate brain WM microstructural alterations of T2DM participants using NODDI and determine its feasibility in detecting global and regional WM differences; and 2) to examine the association between the altered NODDI metrics, the duration of diabetes, and the glycated haemoglobin (HbA1c) levels.

## Materials and Methods

### UK Biobank Cohort

UK Biobank is a population-level study of 500,000 people aged 40 to 69 years old recruited between 2006 and 2010, with data collected in twenty-two centres across England, Scotland, and Wales^22^. All participants provided informed consent forms. Ethical approval was obtained from the Northwest Multi-centre Research Ethics Committee. The current study was approved under the UK Biobank application ID 43822. Details of the UK Biobank cohort and consent are available on the UK Biobank website^22^.

### Clinical Characteristics and Sample Selection

A total of 33,667 participants were included in this study (3338 participants with T2DM and 30329 non-T2DM participants). Touch-screen questionnaires, health exams, brief computer-assisted interviews, biological samples, and imaging data were among the baseline assessments^21^. At the time of the imaging visit, data on age, sex, qualification, diabetes history, body mass index (BMI), blood pressure (BP), glycated haemoglobin (HbA1c), cardiovascular disease history, and neurological disorders were collected and reported. Socioeconomic status, family history, lifestyle, and health status were among the datasets acquired through questionnaires and interviews.

Based on physician diagnoses and summary diagnoses recorded across all UK hospitals, we included participants with T2DM who did not have neurological diseases such as stroke, haemorrhage, cerebral infarction, Parkinson’s disease, Alzheimer’s disease, epilepsy, head injuries, or tumours. Participants in the UK Biobank with no history of diabetes or neurological disorders were used as a control group. The summary diagnosis category contains summary fields relating to diagnoses made during hospital inpatient admissions. Also, the diagnosis data-fields list for each participant and all the distinct values for that data fields in the inpatient data, including primary and secondary diagnoses, were coded according to the International Classification of Diseases (ICD-10) (see supplementary Table 1 for codes and field IDs).

### Brain MRI Acquisition

Participants in the UK Biobank were invited to attend MRI scans at one of three dedicated imaging centres equipped with identical scanners (3T Siemens Skyra, software VD13A via Siemens Healthcare) and using a Siemens 32-channel receive head coil. Diffusion images were acquired using echo-planar with single-shot Stejskal-Tanner pulse sequence in 100 diffusion-weighted directions (b values=0, 1000, and 2000 s/mm^2^). T1-weighted structural brain images were acquired using a three-dimensional MPRAGE sequence. The brain imaging protocol details are available online and in the UK Biobank imaging report^23,24^.

### Diffusion Image Processing

In the diffusion data, the anterior-posterior (AP) encoding direction is corrected for eddy currents and head motion, and outlier-slices (individual slices in the 4D data) are corrected using the eddy tool in the first step of this pipeline^24–26^. This step requires knowledge of the “best” b = 0 image in the AP direction. The primary corrections performed by eddy was performed in-plane. The gradient distortion correction is applied after eddy to produce a more precise correction^27^. The diffusion images were analysed based on tract-skeleton (TBSS) processing. The output was a wide range of diffusion MRI-derived measures within different tract areas: 1) measurements deriving from diffusion-tensor modelling, and 2) measurements deriving from microstructural model fitting. More details on image processing are available in previous UK Biobank reports^20,24,28^.

### DTI and NODDI fitting

The DTI fitting was applied to the b=1000 shell (50 directions) using DTIFIT tool^29^. In addition to DTI fitting, the entire two-shell diffusion data set was fed into NODDI modelling, which was performed with the AMICO (Accelerated Microstructure Imaging via Convex Optimization) tool^30^. This aims to generate voxel-wise microstructural parameters such as ICVF, ODI and IsoVF.

### White matter tract skeleton analysis

The DTI FA image was fed into TBSS^31^, which aligned the FA image onto a standard-space-WM skeleton, with alignment improved over the original TBSS skeleton-projection methodology by employing a high-dimensional FNIRT-based warping technique^32^. The resulting standard-space warp was applied to DTI/NODDI output maps. For each DTI/NODDI map, the skeletonized images were averaged within a set of 48 standard-space tract masks defined by the Johns Hopkins University group (JHU-ICBM atlas)^33^ to generate a set of diffusion imaging-derived phenotypes (IDPs).

### Confound Modelling in UK Biobank Brain Imaging

A huge imaging dataset, such as the UK Biobank data, may contain confounds that must be corrected before running the final statistical analysis. In this study, the following imaging and demographic features were de-confounded from brain imaging-derived phenotypes (IDPs): age, sex, head motion, head size scaling, confounds related to bed/table position in the scanner (for the three axes X, Y, Z), confounds of acquisition date/time, and imaging sites^20,34^. The UK Biobank dataset was de-confounded with a linear model concurrently (i.e., in a single regression-based de-confounding employing all confound variables combined). For instance, Y is the N-vector of interest and X is the N-by-P matrix of all confounds. In that case, the confound-adjusted variables are the residuals from the linear model fit of Y to X^34^.

### Statistical Analysis

Statistical analysis was performed using SPSS 27.0 (IBM, NY, USA). Continuous data were presented as mean ± standard deviation, and categorical data were presented as percentages. Intergroup differences, including clinical characteristics and brain WM differences, were compared by student’s t-test and Pearson’s chi-square test. Pearson correlation analyses were conducted to explore the association between HbA1c, disease duration and diffusion measures. Pearson’s correlation coefficient was considered weak (0< r ≤0.4). The defined *P-value* threshold was set to a *P*-value of 0.05 (false discovery rate).

## Result

### Clinical characteristics

Of the 39,000 individuals participating in the entire UK Biobank imaging cohort, 3,338 participants with T2DM and 30,329 non-T2DM participants had undergone diffusion MRI. They were retained in this analysis after applying the inclusion/exclusion criteria (Fig.1). Characteristics of both groups are presented in Table 1.

**Figure 1.**
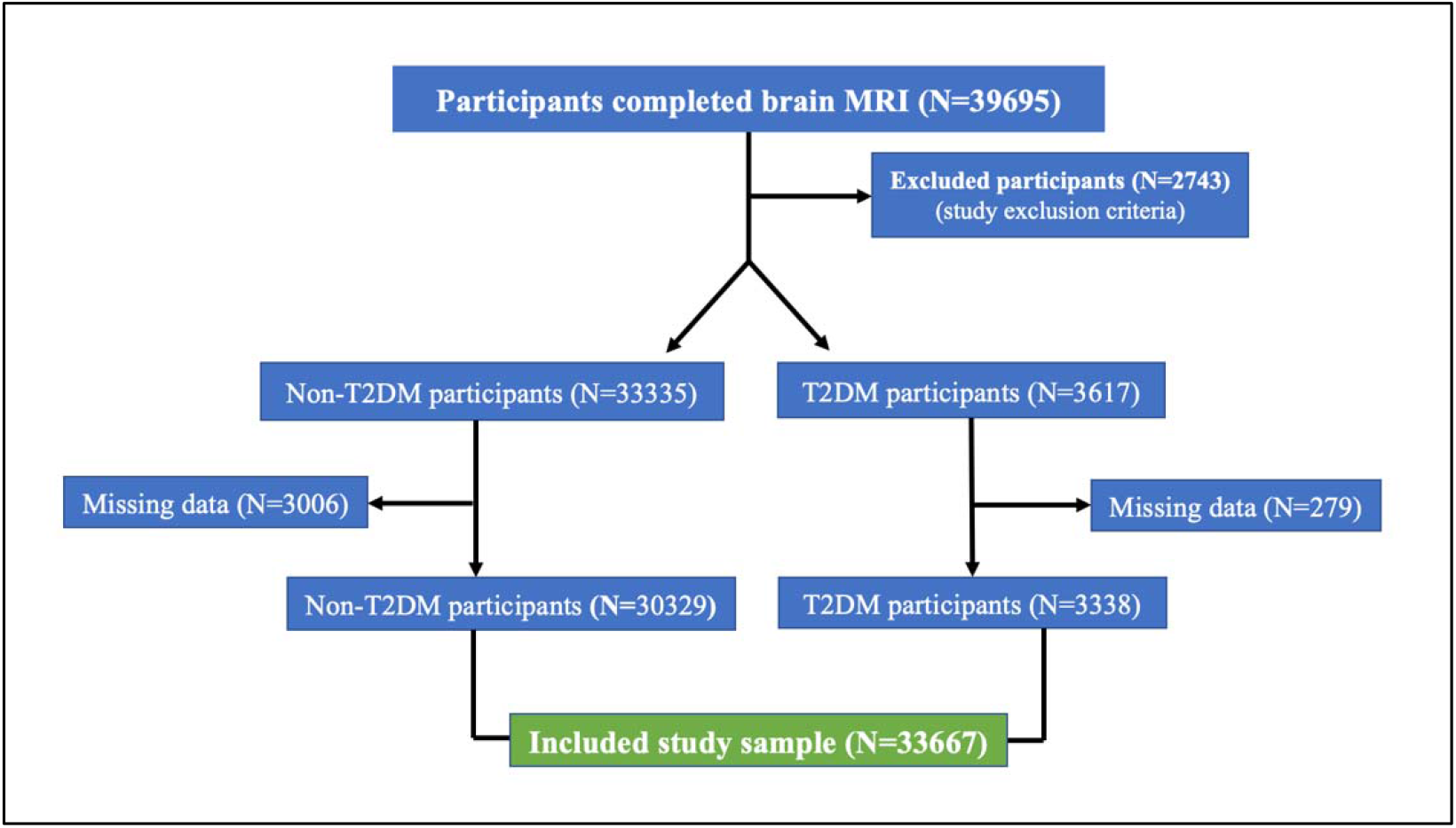
Flowchart of UK Biobank participants included in this study.

**Table 1.**
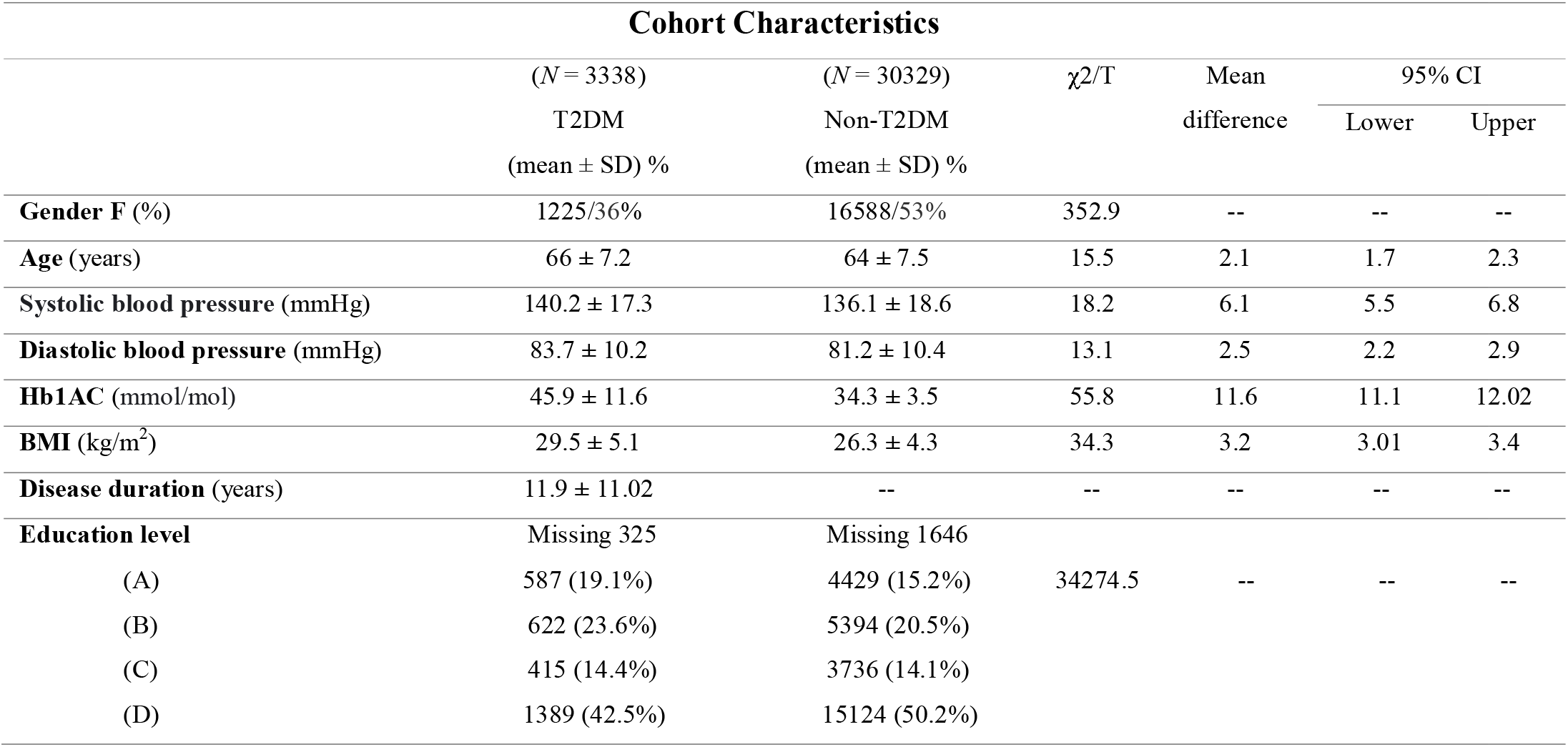
The clinical characteristics of the included participants from the UK Biobank database. ± = mean and standard deviation; A = College or University degree; B = A levels/AS levels or equivalent; C = O levels/GCSEs or equivalent; D = CSEs or equivalent, -- = not applicable.

| Cohort Characteristics |  |  |  |  |  |  |
| --- | --- | --- | --- | --- | --- | --- |
| | (N = 3338)<br>T2DM<br>(mean ± SD) % | (N = 30329)<br>Non-T2DM<br>(mean ± SD) % | $\chi^2/T$ | Mean<br>difference | 95% CI | |
|  |  |  |  |  | Lower | Upper |
| **Gender F (%)** | 1225/36% | 16588/53% | 352.9 | -- | -- | -- |
| **Age (years)** | 66 ± 7.2 | 64 ± 7.5 | 15.5 | 2.1 | 1.7 | 2.3 |
| **Systolic blood pressure (mmHg)** | 140.2 ± 17.3 | 136.1 ± 18.6 | 18.2 | 6.1 | 5.5 | 6.8 |
| **Diastolic blood pressure (mmHg)** | 83.7 ± 10.2 | 81.2 ± 10.4 | 13.1 | 2.5 | 2.2 | 2.9 |
| **Hb1AC (mmol/mol)** | 45.9 ± 11.6 | 34.3 ± 3.5 | 55.8 | 11.6 | 11.1 | 12.02 |
| **BMI (kg/m<sup>2</sup>)** | 29.5 ± 5.1 | 26.3 ± 4.3 | 34.3 | 3.2 | 3.01 | 3.4 |
| **Disease duration (years)** | 11.9 ± 11.02 | -- | -- | -- | -- | -- |
| **Education level** | Missing 325 | Missing 1646 |  |  |  |  |
| (A) | 587 (19.1%) | 4429 (15.2%) | 34274.5 | -- | -- | -- |
| (B) | 622 (23.6%) | 5394 (20.5%) |  |  |  |  |
| (C) | 415 (14.4%) | 3736 (14.1%) |  |  |  |  |
| (D) | 1389 (42.5%) | 15124 (50.2%) |  |  |  |  |

### WM Intergroup Differences and Association with Metabolic Profile: Overview

Parametric maps of DTI and NODDI were generated from each subject. This large sample study revealed subtle, but global WM microstructural changes in the participants with T2DM compared to non-T2DM individuals, which were reflected by reduced FA and ICVF, increased MD, AD, RD, ODI, and IsoVF. These subtle alterations had weak correlations with disease duration and HbA1c.

### DTI Intergroup WM Differences

FA showed a significant reduction in most of the examined WM tracts of participants with T2DM compared to non-T2DM individuals (all *p*<0.05, d < 0.3). Also, MD, AD and RD showed a significant increase in a majority of investigated WM tracts of participants with T2DM (all *p*<0.05, d < 0.3) (see Fig. 2 and supplementary Table 5). Details on statistics of DTI differences are shown in supplementary Table 2A-D. *NODDI Intergroup WM Differences* ICVF was significantly reduced in a majority of the investigated WM tracts of participants with T2DM compared to non-T2DM individuals (all *p*<0.05, d < 0.3). Contrary to the observed ICVF decrease, ICVF values increased in the right cerebral peduncle (*p*<0.001) in T2DM participants. ODI and IsoVF were significantly increased in a majority of investigated WM tracts of participants with T2DM compared to non-T2DM individuals (all *p*<0.05, d < 0.2). In contrast, there was a decrease in ODI in the bilateral posterior limbs of the internal capsules (*p*<0.001) and a reduction of IsoVF in the middle cerebral peduncle, right corticospinal tract, and right cingulum cingulate gyrus (*p*<0.05) in T2DM participants (see Fig.2 and supplementary Table 5). Details on statistics of NODDI differences are shown in supplementary Tables 3A-C.

**Figure 2.**
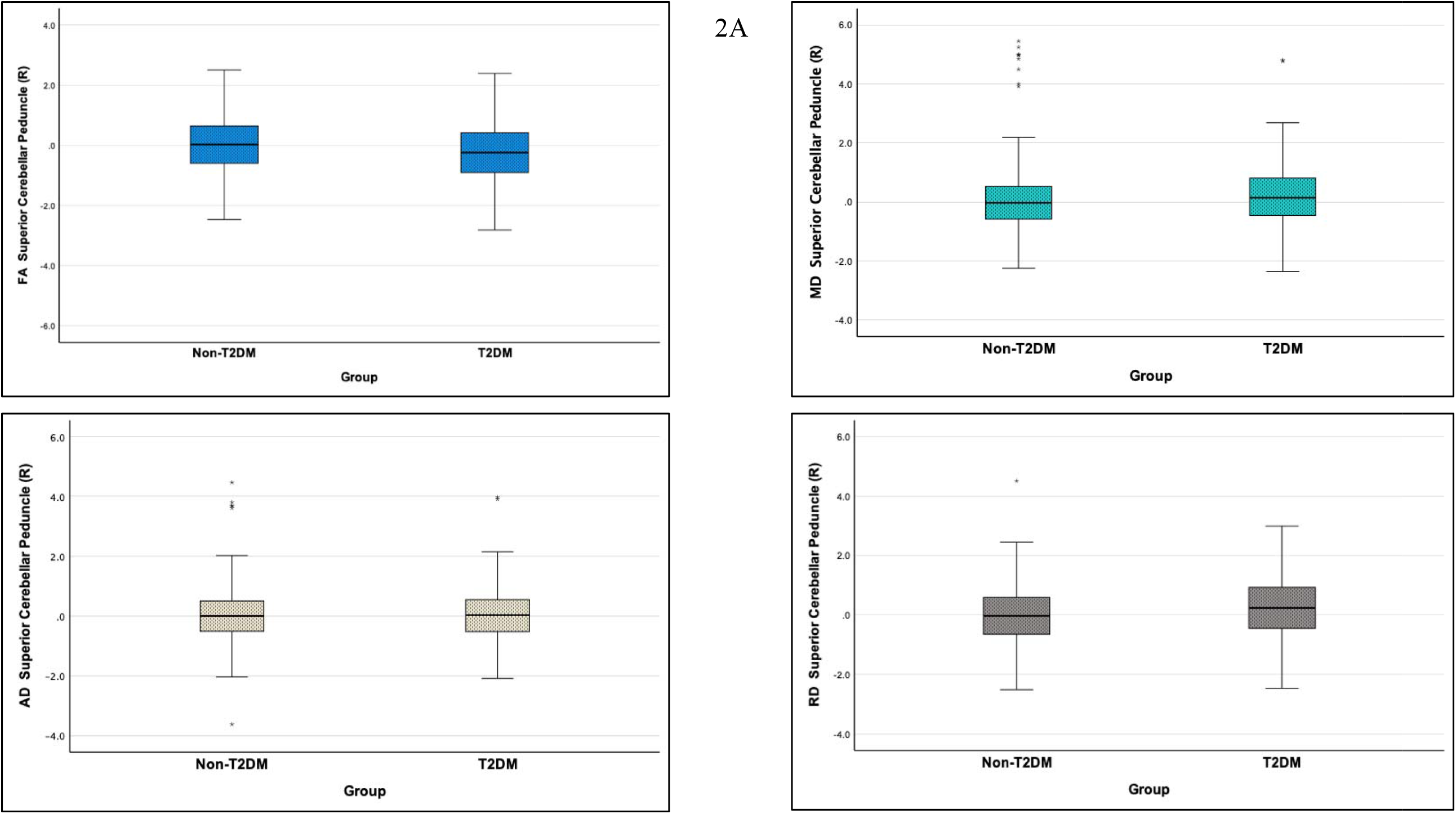
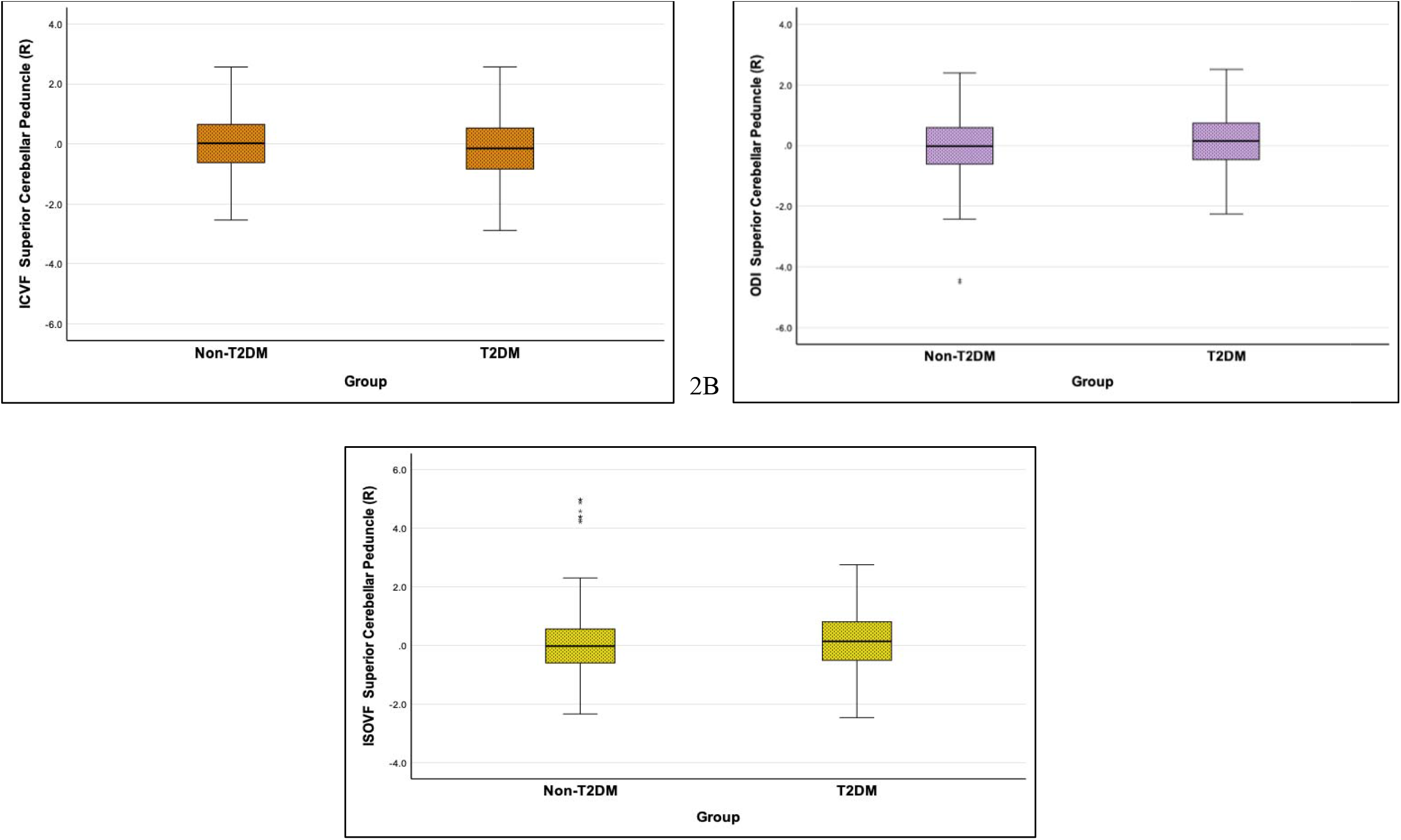
The boxplots for the right SCP as a selected WM tract (from the de-confounded dataset) with a larger effect size to visualise the intergroup WM alteration in patients with T2DM (*P* <0.05, false discovery rate adjustment). 2A. represents SCP changes detected by DTI metrics; 2B. illustrates SCP changes detected by NODDI metrics.

### Diffusion Measures and Correlation with Metabolic Profile

#### Disease duration, HbA1c and DTI

Correlation analysis showed weak association between disease duration, HbA1c, and DTI changes in WM tracts of participants with T2DM. Reduced FA and increased MD, AD and RD correlated with longer disease duration in most examined WM tracts (all *p*<.05) (0< r ≤0.2). Also, reduced FA and increased MD, AD and RD weakly correlated with a higher level of HbA1c (all *p*<.05) (0< r ≤0.2) (see Fig.3, Table 2, and supplementary Table 4).

**Figure 3.**
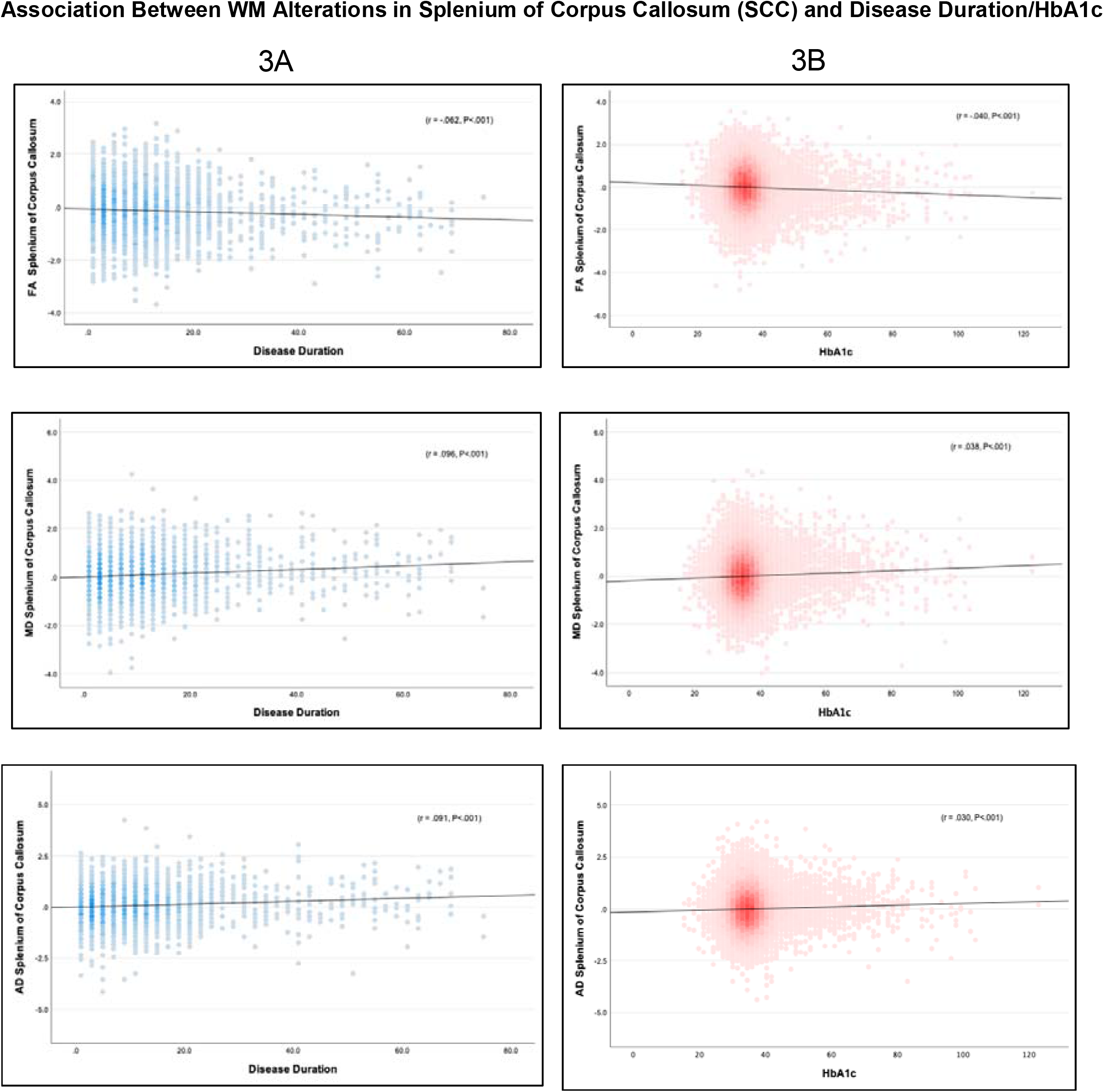
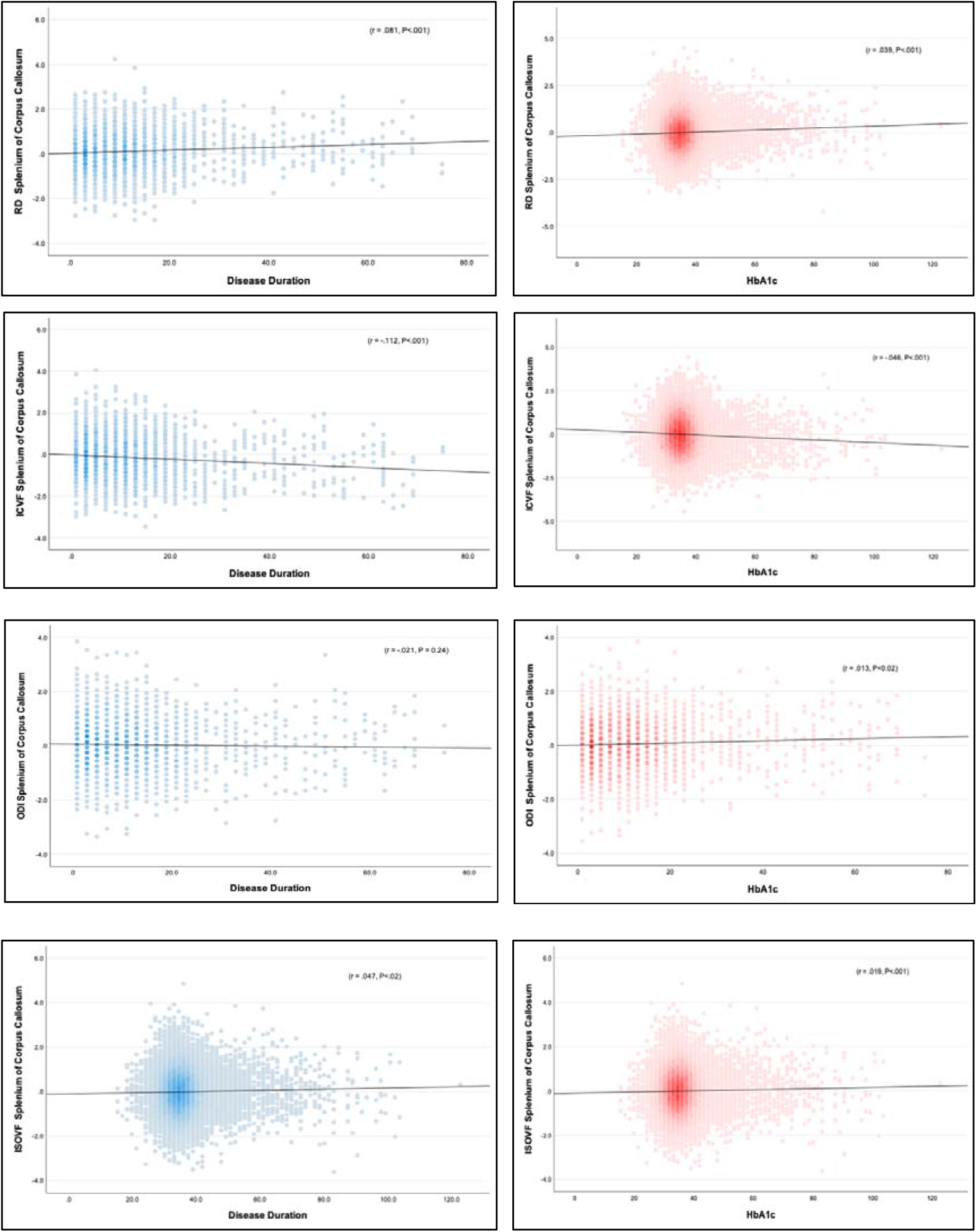
The splenium of corpus callosum (SCC) as a selected WM tract from the de-confounded dataset to visualise the association between the WM change detected by DTI/NODDI and metabolic profile. (2A) illustrates the association between WM alterations in SCC with disease duration; (2B) shows the association of WM alterations in SCC with HbA1c.

**Table 2.**
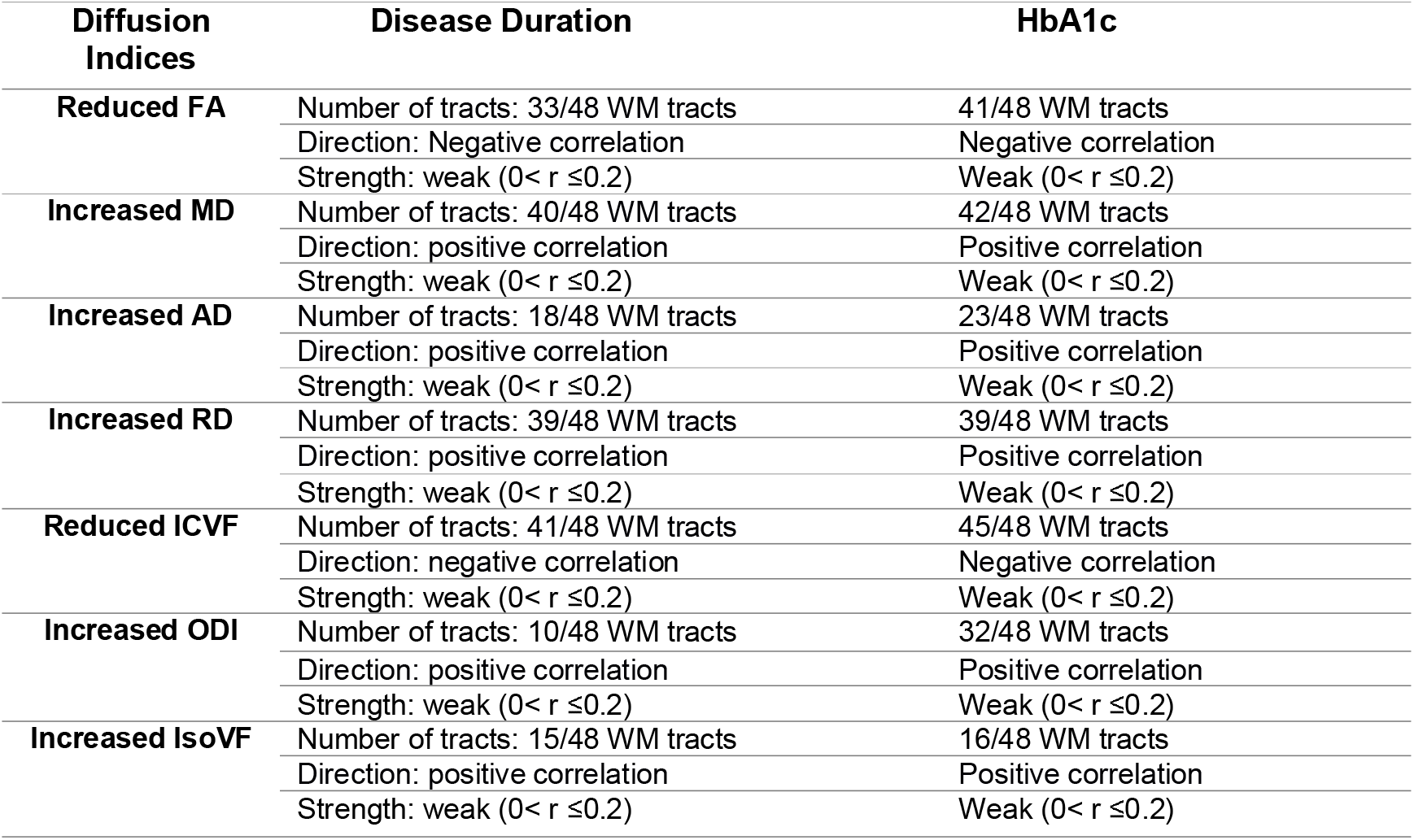
The number of WM tracts that correlated with disease duration and HbA1c levels, direction, and strength of the correlations (*P* <0.05 after false discovery rate adjustment).

| Diffusion Indices | Disease Duration | HbA1c |
| --- | --- | --- |
| **Reduced FA** | Number of tracts: 33/48 WM tracts | 41/48 WM tracts |
|  | Direction: Negative correlation | Negative correlation |
| | Strength: weak ($0 < r \leq 0.2$) | Weak ($0 < r \leq 0.2$) |
| **Increased MD** | Number of tracts: 40/48 WM tracts | 42/48 WM tracts |
|  | Direction: positive correlation | Positive correlation |
| | Strength: weak ($0 < r \leq 0.2$) | Weak ($0 < r \leq 0.2$) |
| **Increased AD** | Number of tracts: 18/48 WM tracts | 23/48 WM tracts |
|  | Direction: positive correlation | Positive correlation |
| | Strength: weak ($0 < r \leq 0.2$) | Weak ($0 < r \leq 0.2$) |
| **Increased RD** | Number of tracts: 39/48 WM tracts | 39/48 WM tracts |
|  | Direction: positive correlation | Positive correlation |
| | Strength: weak ($0 < r \leq 0.2$) | Weak ($0 < r \leq 0.2$) |
| **Reduced ICVF** | Number of tracts: 41/48 WM tracts | 45/48 WM tracts |
|  | Direction: negative correlation | Negative correlation |
| | Strength: weak ($0 < r \leq 0.2$) | Weak ($0 < r \leq 0.2$) |
| **Increased ODI** | Number of tracts: 10/48 WM tracts | 32/48 WM tracts |
|  | Direction: positive correlation | Positive correlation |
| | Strength: weak ($0 < r \leq 0.2$) | Weak ($0 < r \leq 0.2$) |
| **Increased IsoVF** | Number of tracts: 15/48 WM tracts | 16/48 WM tracts |
|  | Direction: positive correlation | Positive correlation |
| | Strength: weak ($0 < r \leq 0.2$) | Weak ($0 < r \leq 0.2$) |

#### Disease duration, HbA1c and NODDI

Reduced ICVF and increased ODI and IsoVF had weak correlations with longer disease duration and HbA1c in most examined WM tracts (all *p*<.05) (0< r ≤0.2). Also, reduced ICVF and increased ODI and IsoVF had a weak but significant correlation with higher levels of HbA1c (all *p*<.05) (0< r ≤0.2) (see Fig.3, Fig. 4, Table 2, and supplementary Table 4).

**Figure 4.**
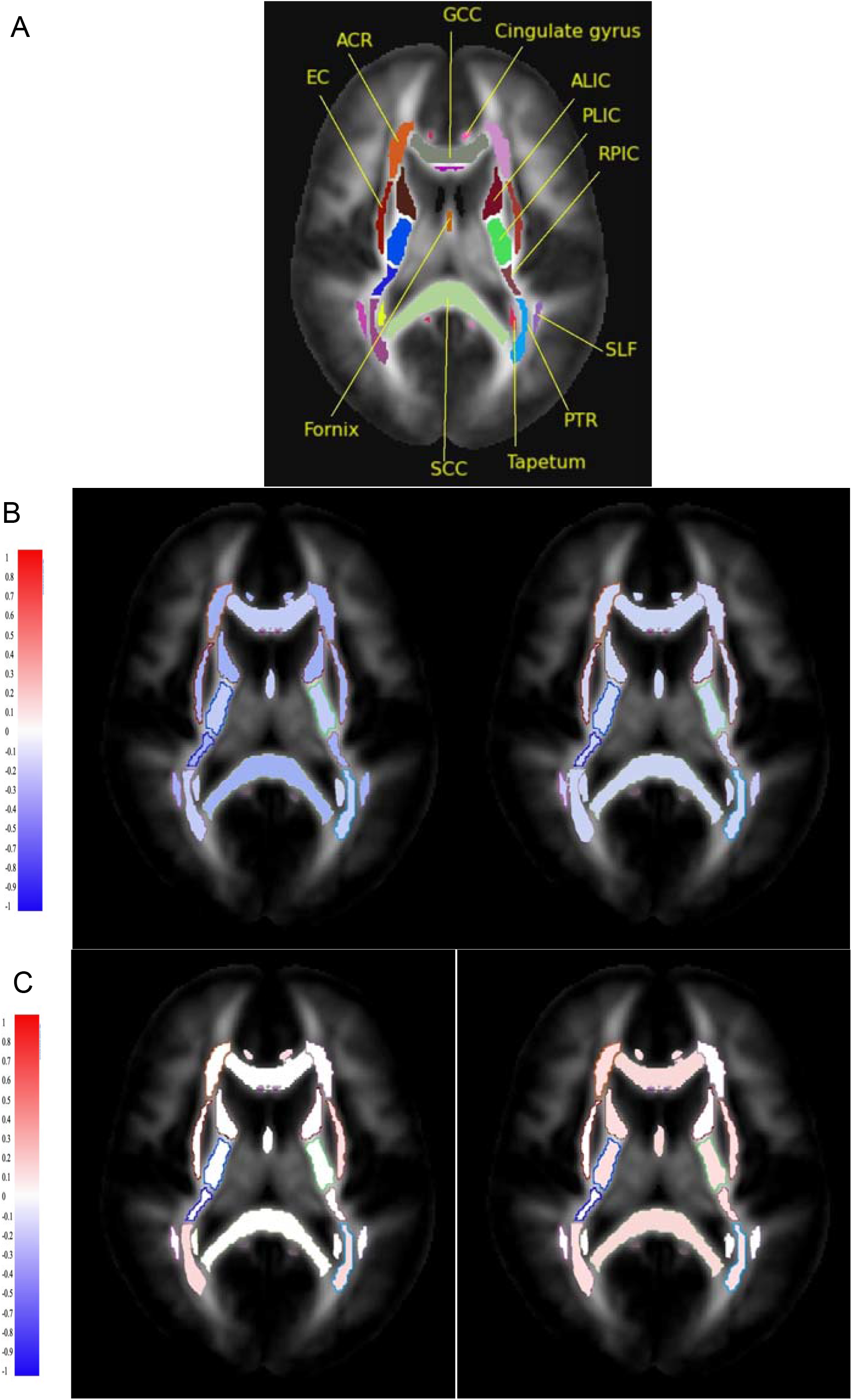
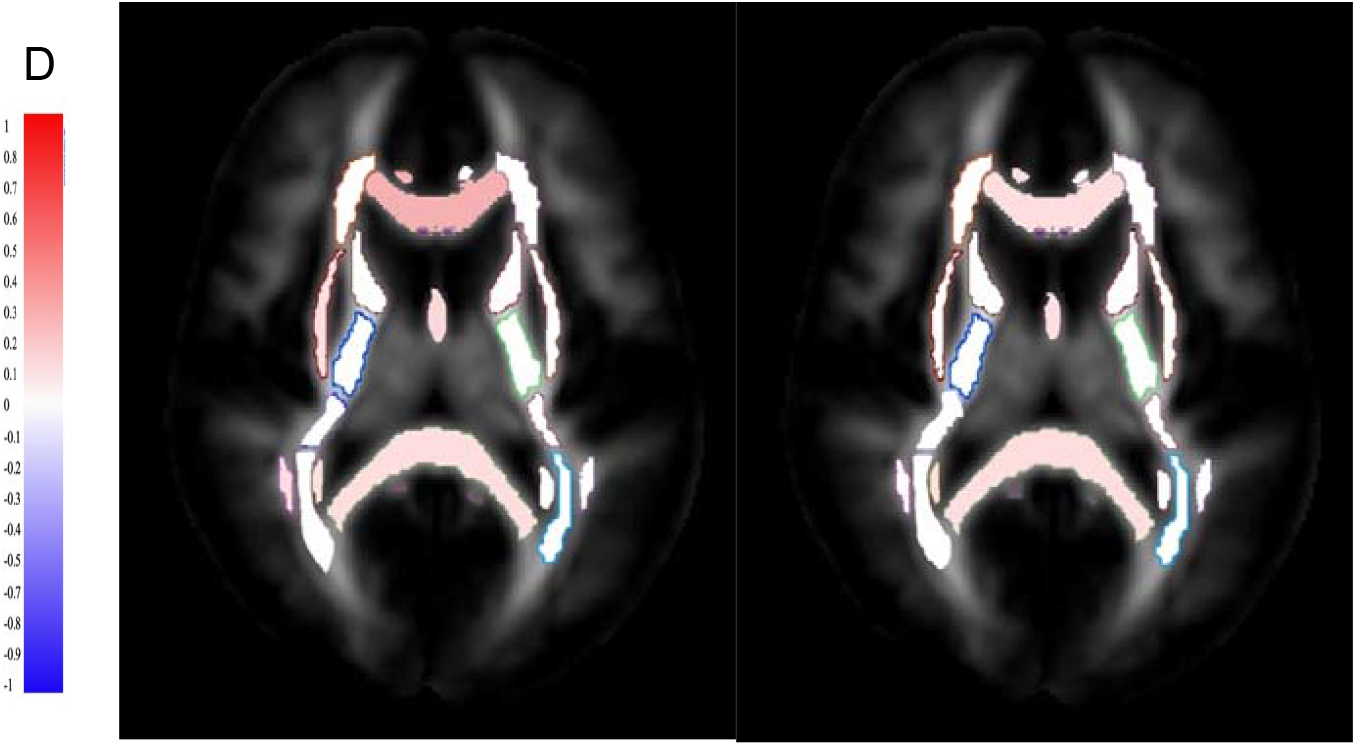
The correlations between disease duration and HbA1c with major WM tracts in participants with T2DM, and these WM structures are only selected for visualising purposes. (A) WM tracts included: Genu of corpus callosum (GCC), fornix, cingulate of gyrus, superior longitudinal fasciculus (SLF), anterior corona radiata (ACR), anterior limb of internal capsule (ALIC), posterior limb of internal capsule (PLIC), posterior thalamic radiation (PTR), tapetum, splenium of corpus callosum (SCC), external capsule (EC), and retrolenticular part of internal capsule (RPIC). (B) Altered ICVF and disease duration/HbA1c. (C) Altered ODI and disease duration/HbA1c. (D) Altered IsoVF and disease duration/HbA1c. Red = positive correlation; blue = negative correlation.

## Discussion

Using this large-scale diffusion imaging data from 33,667 participants from the UK Biobank, we evaluated the ability of DTI and NODDI-derived metrics to capture microstructural changes in the WM in participants with T2DM. Moreover, we examined whether disease duration and HbA1c are associated with altered DTI/NODDI parameters.

In T2DM participants, WM tracts and fibres mostly show reduced FA and ICVF, and increased MD, AD, RD, ODI, and IsoVF, as presented in supplementary Table 5. Reduced anisotropy and increased diffusivity reflected by reduced FA and higher MD confirm widespread microstructural alterations and broadly agree with previous DTI studies (see the previous reviews^4,10,11^). Moreover, the results revealed that the elevation of AD and RD in participants with T2DM might lead to changes in FA and MD, as AD is linked to axonal integrity and RD is linked to myelin abnormalities. Therefore, the findings of our UK Biobank study suggest that increased interstitial space between the myelin-covered axons (expansion of extracellular space due to the degeneration of fibres/axonal injury) and reduced myelin integrity play a role in WM deficits in T2DM participents^14,35–37^.

The reduction of ICVF in the WM of T2DM in this study was consistent with a previous study^19^, which indicates reduced neurite density. The reduced ICVF or neurite density in T2DM is probably relevant to structural morphology changes such as axonal loss, axonal damage, or axonal demyelination/degeneration^38,39^. Furthermore, the decreased neurite density in participants with T2DM may be accompanied by axonal disorganization, myelin loss, and loss of fibre orientation coherence that may lead to increased ODI in response to alterations in WM^10,40–42^. On the contrary, increased ICVF in a few WM tracts (e.g., right cerebral peduncle and left tapetum) may indicate an early cellular swelling such as astrocyte/axonal swelling or activation of adaptive axons as they correlate with shorter disease duration and lower HbA1c levels^43–46^. Interestingly, the result shows decreased ODI is observed in a few WM tracts, such as bilateral posterior limbs of internal capsules, which may reflect the loss of neuronal/crossing fibres, reorganisation of fibres, or reduced axonal dispersion in T2DM^39,47,48^. In addition, higher coherence of axonal packing in the WM of T2DM patients reflected by the decrease of ODI may suggest either a reduced collateral branching driven by the pathology or alterations in the morphology of an individual axon. Lastly, the elevated IsoVF may indicate vasogenic edema or increased extracellular free water, which may be caused by the disruption of blood-brain barriers (BBB) in participants with T2DM^44^, or may indicate a degree of atrophy by reflecting cell shrinkage and decreased tissue volume fraction^49^.

By examining the relationship between the metabolic profile and WM intergroup differences, we identified statistically significant associations between altered DTI and NODDI parameters in T2DM participants and glycaemic control/disease duration. The neurite density in the WM of T2DM participants reflected by a reduced ICVF has a consistent association with disease duration and poorly-controlled blood glucose, suggesting that T2DM participants with longer disease duration and higher plasma glucose tend to have reduced neurite density. On the other hand, increased ICVF values in the right cerebral peduncle and left tapetum are associated with shorter disease duration, suggesting that ICVF may detect neurite deterioration at early disease stages. For ODI, poorly-controlled blood glucose and disease duration correlate with loss of coherence of WM fibres or axonal disorganisation reflected by higher ODI. However, the ODI association with the metabolic profile was not as consistent as the ICVF correlation, particularly with disease duration. Also, reduced ODI in the posterior limb of the internal capsule shows an association with disease duration, indicating the loss of neuronal fibres or neurodegeneration may occur with a longer disease duration. The vasogenic edema or increased interstitial fluids reflected by increased IsoVF in T2DM participants is associated with a longer disease duration and higher HbA1c levels. Regarding age and disease duration, it is worth noting that we tested two models with and without age adjustment, and the results suggest that age does not affect the association between WM alterations in T2DM participants and disease duration.

Although the findings of this study demonstrate that DTI still has a potential value as a clinical biomarker in T2DM, there are acknowledged challenges in relating standard DTI parameters directly to pathological or biological changes. This study may show that the changes captured by NODDI parameters may contribute to FA alterations (especially in areas with complex mircostrcutre^50^) in the brain WM of T2DM participants. However, NODDI may suffer the constant diffusivity assumption when pathology changes diffusivity. NODDI model relies on assumptions to speculate the microstructural alterations; nevertheless, the NODDI parameters with different neurological disorders must be interpreted carefully^51–53^.

Our results show that NODDI parameters could detect WM neuropathological changes in WM microstructure, suggesting its potential role for monitoring microstructural alterations in T2DM. Nevertheless, this study has limitations. It is retrospective and based on a biobank rather than a prospective case-control study. Also, there could have been other non-excluded systemic conditions that could impact the NODDI metrics, such as previous chemotherapy, cardiac surgery, chronic diseases, and psychiatric disorders. Further work to improve this novel study is suggested as follows: the impact of T2DM on brain functions and neurocognition and their association with the altered NODDI parameters was not explored. Also, the study did not include a follow-up assessment to monitor the WM changes longitudinally in T2DM participants. Lastly, the microstructural impact of T2DM on the grey matter is not evaluated in this study.

## Conclusion

WM microstructural abnormalities in T2DM participants are associated with longer disease duration and higher glycated haemoglobin, reflecting poorer glucose control. As an advanced diffusion model, NODDI can characterize WM neuroaxonal pathology in T2DM, giving quantitative biophysical information for understanding the impact of T2DM on brain WM microstructure.

## Data Availability

All data produced in the present study are available upon reasonable request to the authors.

## Author Contributions

Ali-Reza Mohammadi-Nejad and Stamatios N. Sotiropoulos assisted with providing data and methods from the UK Biobank and content review. Chris Tench, Anna Podlasek, and Amjad Altokhis helped with the analysis steps. Sieun Lee, Cris Constantinescu and Rob Dineen supervised the entire research project.

## Acknowledgements

Data were provided by the UK Biobank under Project ID 43822 (PI: Sotiropoulos). This study was partially supported by the NIHR-funded Nottingham Biomedical Research Centre. Also, this work is supported in part by Nottingham Hospitals Charity (APP 1666/N0185) and King Saud bin Abdul-Aziz University for Health Sciences in Saudi Arabia.

